# The Incidence, Predictors, and Mortality Impact of Ventricular Pacing Dependency after Transcatheter Aortic Valve Replacement: A Prospective Cohort

**DOI:** 10.1101/2022.09.12.22279879

**Authors:** Sirin Apiyasawat, Mann Chandavimol, Natcha Soontornmanokati, Chulaporn Sirikhamkorn

## Abstract

**Background:** Atrioventricular conduction disturbance occurs in a significant number of patients underwent transcatheter aortic valve replacement (TAVR). However, not all of the cases persistently depend on ventricular pacing.

**Objective:** To study the incidence, predictors, and outcomes of new ventricular pacing dependency (VpDep) after TAVR.

**Design:** Prospective, single-center, cohort trial

**Methods:** The first 130 consecutive transfemoral TAVR cases, except 3 with prior cardiac electronic implantable devices (CIEDs) who depended on ventricular pacing, were analyzed. The endpoints were VpDep at 1 month and all-cause mortality at the end of follow-up in 2021. The effects of variables on VpDep and all-cause mortality were evaluated using multivariate binary logistic regression and Cox-regression analysis, respectively. First degree atrioventricular block (AVB) was considered severe when PR interval > 300 ms.

**Results:** Out of 127 patients in the analysis (mean age 82 years, SD 6.3; 62.2% female sex; 67.7% Balloon-expandable device), 7 patients (5.5%) had implanted with CIEDs before TAVR but not ventricular pacing dependent. The incidence of VpDep at 1 month was 7.9% (n=10) among all patients and 34.5% among patients with CIEDs (n=29). VpDep was likely to occur in patients with pre-existing right bundle branch block (OR 21.38; 95% CI 3.25-139.33; P=.001) and severe 1^st^ degree or Mobitz I AVB (OR 14.79; 95% CI 1.65-132.74; P=.016). After a mean follow-up of 25.8 months (SD 21.2), death from any causes occurred in 18 patients (14.2%). VpDep was not associated with excess mortality.

**Conclusion:** VpDep at 1 month occurred in 7.9% of all patients underwent TAVR. Pre-existing conduction abnormalities were significantly associated with higher risk of VpDep. Mortality was similar between patients with and without VpDep.

**Clinical Trial Registration:** Thai Clinical Trial Registration; Study ID: TCTR20220726005

## INTRODUCTION

Atrioventricular conduction disturbance is a common consequence of transcatheter aortic valve replacement (TAVR). Many of these cases require permanent pacemaker (PPM) implantation.^1 2^ However, conduction disturbance may resolve over time and ventricular pacing dependent (VpDep) may occur only in a certain proportion of patients with PPM.^4 4 5^ VpDep could lead ventricular dysfunction^6^ and affect the long-term prognosis of patients underwent TAVR. Prior literatures calculated the incidence of VpDep based on variable selection criteria.^5^ Almost all of them excluded patients who already had PPM even though they were not VpDep. In the present trial, we studied a cohort of patients underwent TAVR using all types of TAVR devices. The incidence, predictor, and outcome of new VpDep in patients with and without prior PPM were analyzed.

## METHODS

<u>RA</u>mathibodi trans<u>C</u>atheter aortic valve replacement <u>R</u>egistry (RACR) consecutively collected data on all transcatheter aortic valve replacement performed in a tertiary care cardiac center in Thailand. The registry was designed to provide short and long-term clinical outcomes of patients treated with government-approved TAVR devices. Patients with severe symptomatic aortic stenosis were screened and selected by the multidisciplinary heart team using clinical and anatomical imaging information.

In this analysis, the first 130 consecutive transfemoral TAVR cases were studied for the pre-specified endpoints of VpDep at 1 month and all-cause mortality at the end of follow-up in 2021. Patients who already implanted with cardiac implantable electronic device (CIED) and depended on ventricular pacing were excluded from the analysis (Figure 1). The study was approved by the local human research ethics committee.

**Figure 1.**
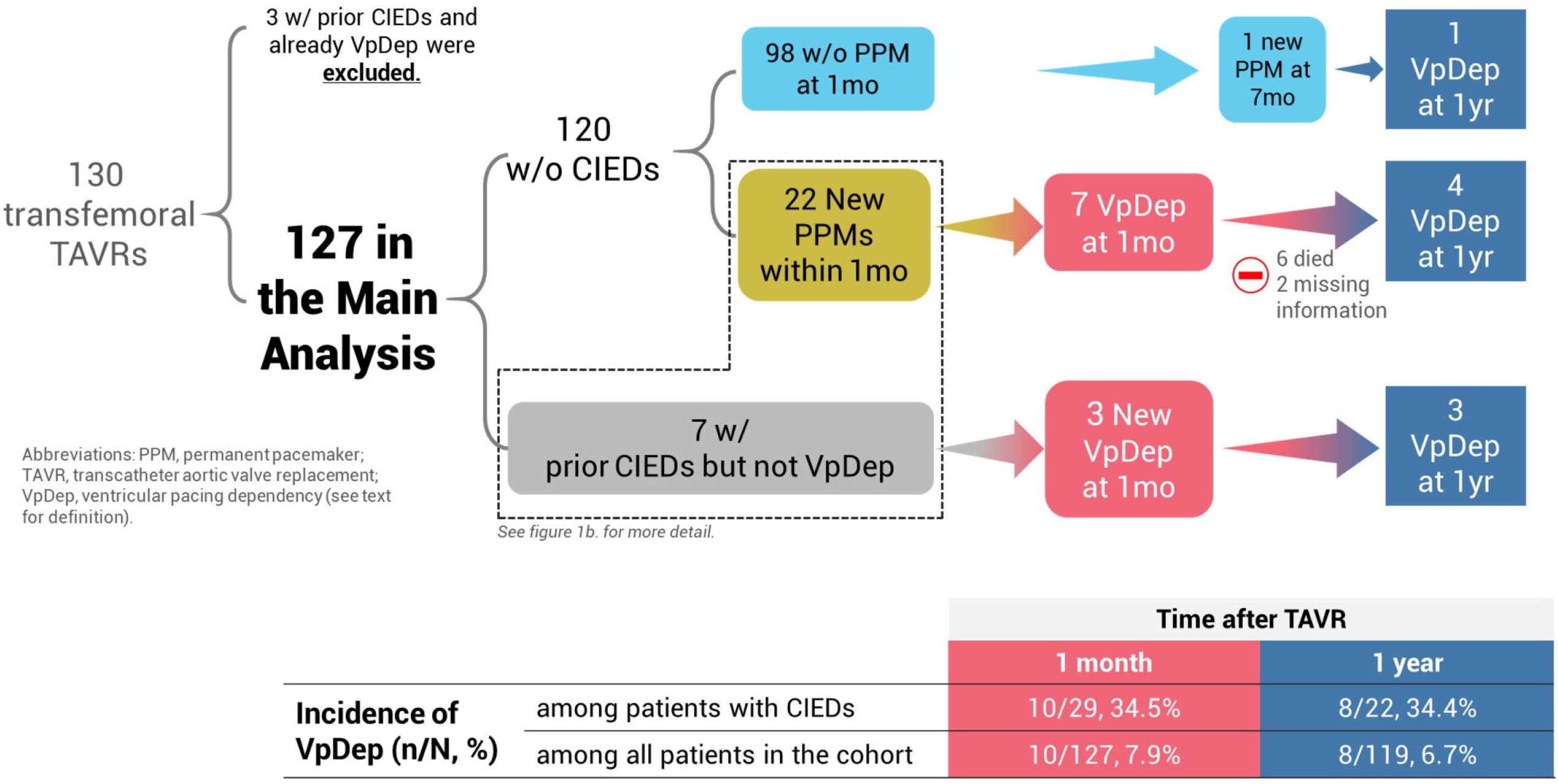
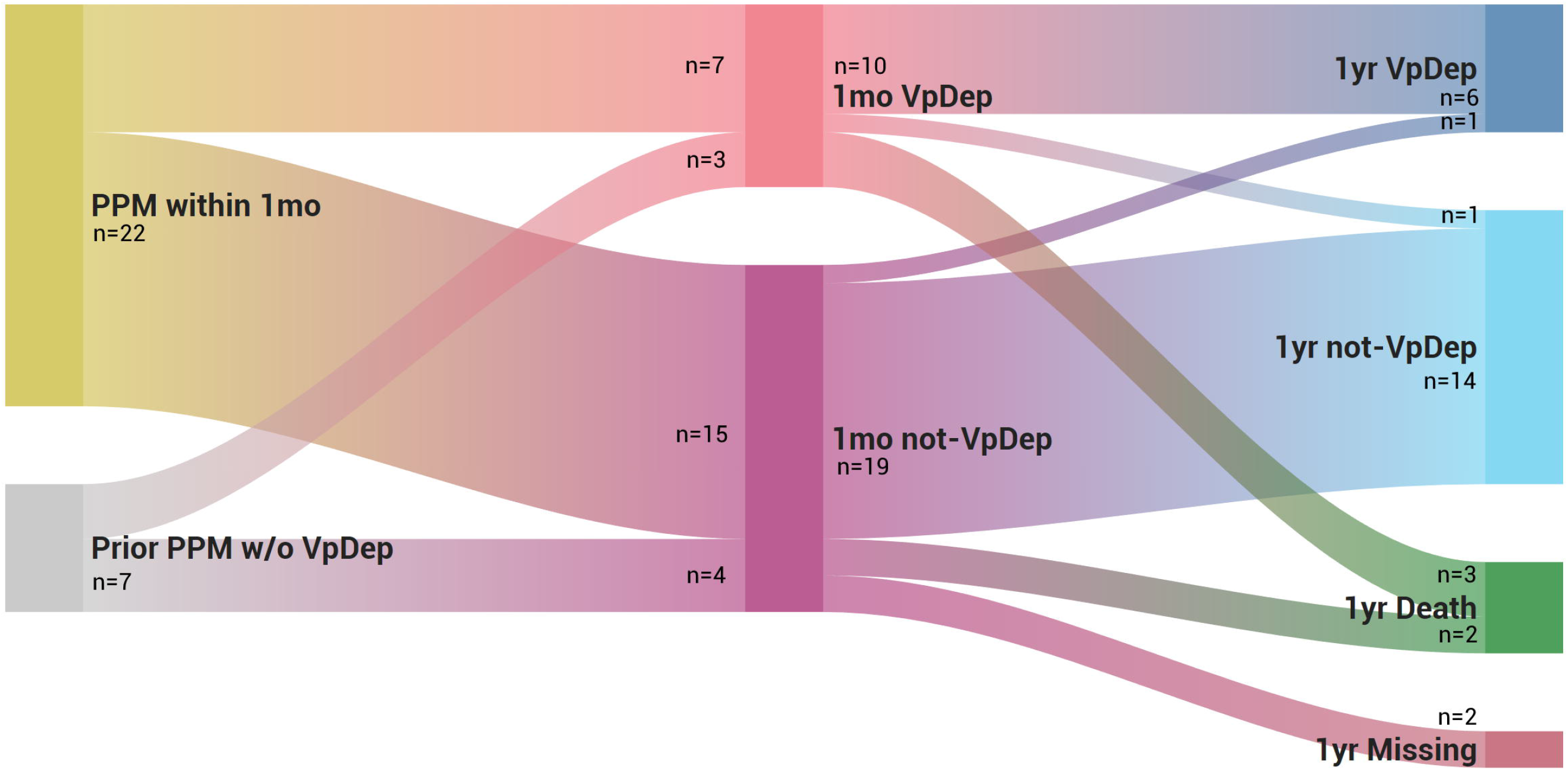
Study flow, incidence of ventricular pacing dependency, and changes in dependency status over time. Figure 1a. Pacemaker status and dependency of all patients. Figure 1b. Changes in ventricular pacing dependency status between 1 month and 1 year among patients implanted with PPM within 1 month or before TAVR (n=29).

### Procedures and TAVR Devices

Transesophageal echocardiography (TEE) and/or computed tomography (CT) were used for anatomical guide and aortic annulus sizing. The device selection and the choices of sedation were left to the discretion of the heart team. The procedure was performed in the cardiac catheterization laboratory or hybrid operating room. After the procedure, all patients were monitored with continuous ECG for at least 72 hours.

Implantation depth was assessed angiographically after the deployment of the device. A total of 10 ml of contrast agent was injected to assess the position of the prostheses. The maximal distance between the intraventricular end of the prosthesis and the aortic annulus at the level of each of all 3 cusps was measured using the HeartVision 2 system (GE Medical Systems SCS, France). The measurement was performed by an intervention cardiologist who was blinded to the results.

### Data Collection

Data, including patient characteristics, valvular parameters, procedural data, and clinical outcomes, were collected in a dedicated database. All patients received a follow-up at 30 days for clinical improvement, major adverse cardiovascular events (MACEs), and another follow-up during the year 2021 for all-cause mortality. For those with CIEDs, data on device settings and VpDep were recorded at the index procedure (if implanted before), 1 month, and 1 year after the index procedure. All follow-ups were performed on the basis of clinical visits or phone calls.

### Definitions

Major bleeding, major vascular complications, and acute kidney injury were defined according to the standardized endpoints by the Valve Academic Research Consortium-2 consensus.^7^ Procedural related conduction disturbances included new or worsening atrioventricular (AV) conduction required electrophysiologic study (EPS) or permanent pacemaker (PPM) implantation.

MACEs at 30 days consisted of death, stroke, myocardial infarction, valve dysfunction, hospitalization for valve-related symptoms or worsening HF, and need for cardiovascular intervention.

Severe 1^st^ degree atrioventricular block (AVB) was diagnosed in a case of 1^st^ degree AVB with PR interval > 300ms.

VpDep was defined as the occurrence of symptoms and signs that create emergent or urgent clinical situation upon abrupt cessation of pacing or the absence of intrinsic ventricular rate of more than 30 bpm.^8^ New VpDep included patients with newly implanted CIEDs within 1 month of the index procedure and became VpDep at 1 month after. Among patients with prior CIEDs, those who were not VpDep before but developed VpDep 1 month after the procedure was also considered to have new VpDep. Evaluation for VpDep was performed in pacemaker clinic by either cardiac electrophysiologist or cardiac device specialist.

### EPS and PPM

Persistent high-grade AV block after 48 hours was the primary indication for PPM implantation. In patients who developed new onset or worsening left bundle branch block (LBBB) that persisted beyond 48 hours, we performed EPS and prophylactically implanted PPM for HV interval ≥65ms.

Types of CIEDs were selected according to the standard guidelines.^9^ Briefly, single-chamber PPM was selected for patients with chronic atrial fibrillation (AF), cardiac resynchronization therapy (CRT) for those who need frequent ventricular pacing with left ventricular systolic function (LVEF) <50%, and dual-chamber PPM for those without above conditions.

Pacemaker programming was tailored to each patient’s specific condition and left at the discretion of the electrophysiology team. Atrioventricular delay (AVD) was extended to allow intrinsic ventricular conduction but not too excessive to avoid hemodynamic disadvantage. Algorithm to reduce unnecessary ventricular pacing may be used as appropriate. In patients with chronotropic incompetence, rate responsive was turned on.

### Statistical Analysis

All analyses were generated using SAS software, version 9.04.01 of the SAS OnDemand for Academics (SAS Institute Inc., Cary, NC, USA.). Categorical variables are expressed as numbers and percentages, and continuous variables are expressed as means and standard deviations (SDs). Fisher’s exact test and one-way analyses of variance were used to compare differences in baseline characteristics.

Predictors of new VpDep were analyzed using multivariate binary logistic analysis. Variables included in the model were age, sex, and variables that had significance level of <.01 in the univariate analyses.

The effect of variables on all-cause mortality was estimated using Cox proportional hazard model. Hazard ratios (HRs) with 95% confidence interval (CI) were calculated. The Kaplan-Meier survival curves were plotted and compared using the log rank test. The proportional assumption was validated using Schoenfeld residuals test. P value of <.05 was considered statistically significant.

## Results

Of the total of 130 consecutive patients underwent transfemoral TAVRs, 3 patients were excluded for the reason that they had CIEDs implanted before the procedure and were characterized as VpDep. Thus, 127 patients remained in the analysis (Figure 1a).

The study population (Table 1 and Supplementary Table S1) was mainly elderly (age 82 years, SD 6.3) female (n=79, 62.2%) with intermediate STS mortality score (6.1%, SD 4.5). CIEDs were already implanted in 7 patients (5.5%). Preexisting conduction abnormalities were recorded as followed: 10 (7.9%) right bundle branch block (RBBB), and 7 (5.5%) severe 1^st^ degree or Mobitz I AVB (Sev 1^st^/Mobitz I).

**Table 1.** Comparisons between patients with 30-day ventricular pacing dependency and independency (N=127)

| Characters | Total<br>(N=127) | VpDep at 1 month |  | *P* |
| --- | --- | --- | --- | --- |
|  |  | independent<br>(n=117) | Dependent<br>(n=10) |  |
| Age, mean (SD), y | 81.8 (6.3) | 81.7 (6.1) | 83.7 (8.0) | .075 |
| Female, n (%) | 79 (62.2%) | 75 (64.1%) | 4 (40.0%) | .131 |
| LVEF, mean (SD), % | 62.3 (13.7) | 62.8 (13.4) | 56.3 (15.6) | .125 |
| AVA, mean (SD), cm<sup>2</sup> | 0.7 (0.20) | 0.7 (0.20) | 0.7 (0.18) | .535 |
| MPG, mean (SD), mmHg | 48.9 (11.9) | 49.6 (11.85) | 40.1 (8.91) | .025 |
| STS mortality score, mean (SD), % | 6.1 (4.5) | 6.2 (4.7) | 4.7 (1.7) | .722 |
| LVOT Calcification, n (%) | 15 (11.8%) | 15 (16.9%) | 0 (0.0%) | .273 |
| Bicuspid valve, n (%) | 3 (2.4%) | 3 (2.6%) | 0 (0.0%) | .608 |

| Characters | Total | VpDep at 1 month |  |  |
| --- | --- | --- | --- | --- |
| Pre-existing severe 1<sup>st</sup> degree or Mobitz I AVB, n (%) | 7 (5.5%) | 4 (3.4%) | 3 (30.0%) | <.001 |
| Pre-existing right bundle branch block, n (%) | 10 (7.9%) | 6 (5.1%) | 4 (40.0%) | <.001 |
| Valve type, n (%) |  |  |  | .587 |
| • Balloon expandable | 86 (67.7%) | 80 (68.4%) | 6 (60.0%) |  |
| • Self-expanding | 41 (32.3%) | 37 (31.6%) | 4 (40.0%) |  |
| Implantation depth |  |  |  |  |
| • At NCC, mean (SD), mm | 3.9 (2.45) | 3.9 (2.49) | 4.0 (2.04) | .419 |
| • At RCC, mean (SD), mm | 4.8 (2.54) | 4.8 (2.61) | 5.1 (1.58) | .169 |
| • At LCC, mean (SD), mm | 4.2 (2.73) | 4.7 (2.76) | 5.4 (2.35) | .206 |
| • Mean (SD), mm | 4.5 (2.48) | 4.5 (2.54) | 4.8 (1.85) | .664 |
| HV interval |  |  |  | .073 |
| • Numbers of patients with data, n (%) | 13 (10.2%) | 11 (9.4%) | 2 (20.0%) |  |
| • Mean (SD), ms | 62.9 (17.7) | 61.3 (18.9) | 72.0 (0.0) |  |
| Indication for PPM implantation after TAVR, n (%) |  |  |  | .755 |
| • Complete AVB | 18 (72.0%) | 12 (10.3%) | 6 (60.0%) |  |
| • New LBBB and HV interval ≥ 65ms | 5 (20.0%) | 4 (3.4%) | 1 (10.0%) | |
| • High grade AVB and HV interval ≥ 65ms | 1 (4.0%) | 1 (0.9%) | 0 (0.0%) | |
| • Sick sinus syndrome | 1 (4.0%) | 1 (0.9%) | 0 (0.0%) |  |
| Ventricular pacing at 30 days, mean (SD), % | 40.9 (22.8) | 12.2 (15.9) | 95.6 (7.2) | <.001 |
| Ventricular pacing at 1 year, mean (SD), % | 46.9 (44.1) | 31.1 (36.8) | 85.3 (37.6) | .004 |
| Death, n (%) | 18 (14.2%) | 16 (13.7%) | 2 (20.0%) | .633 |

Abbreviations: AVA, aortic valve area; AVB, atrioventricular block; LBBB, left bundle branch block; LCC, left coronary cusp; LVEF, left ventricular ejection fraction; LVOT, left ventricular outflow tract; MPG, mean peak gradient; NCC, non-coronary cusp; PPM, permanent pacemaker; RCC, right coronary cusp; STS, Society of Thoracic Surgeons.

TAVR was successfully performed in 126 patients (99.2%). One patient (0.8%) suffered from annulus rupture and died during the procedure. Periprocedural stroke, cardiac tamponade, and major bleeding occurred 2 (1.6%), 4 (3.1%), and 4(3.1%) patients respectively (Supplementary Table S2). The implantation depths at non-coronary cusp, right coronary cusp, and left coronary cusp were 3.9 mm (SD 2.45), 4.8 mm (SD 2.54), and 4.7 mm (SD 2.73) respectively (Table 1).

### New Conduction Abnormalities and VpDep

New conduction abnormalities occurred in 37 patients (29.1%), complete AV block being the most common (n=13, 10.2%). A total of 25 patients (19.7%) received PPM implantation after the procedure. Most of them (n=22, 17.3%) were implanted within the first 7 days (Supplementary Figure S1). Of those, 7 patients (31.8%) were classified as VpDep at 1 month after the procedure (Figure 1a). Among patients who had CIEDs implanted prior to the procedure (n=7), 3 patients (42.9%) became VpDep at 1-month period. Therefore, new 1-month VpDep occurred in a total of 10 patients or 7.9% of all patients in the cohort. The rates of VpDep, calculated based on the total numbers of patients with CIEDs, were 34.5% at 1 month and 34.4% at 1 year. At 1 year, most patients (n/N, 20/22; 90.9%) remained in the same VpDep status as they were at 1 month (Figure 1b). The indication, types of pacing devices, and pacemaker settings of all patients with CIEDs are shown in Supplementary Table S3 and Table S4.

Patients with new VpDep at 1 month (Table 1) were more likely to have pre-existing RBBB (n=4, 40%) than those without new VpDep (n=6, 5.1%; P<.001). They are also more likely to have pre-existing Sev 1^st^/Mobitz I (n=3, 30%) than those without new VpDep (n=4, 3.4%; P<.001). In a multivariate analysis, preexisting RBBB (OR 21.38; 95% CI 3.25-139.33; P=.001) and Sev 1^st^/Mobitz I (OR 14.79; 95% CI 1.65-132.74; P=.016) were independently associated with the occurrence of new VpDep (Figure 2).

**Figure 2.**
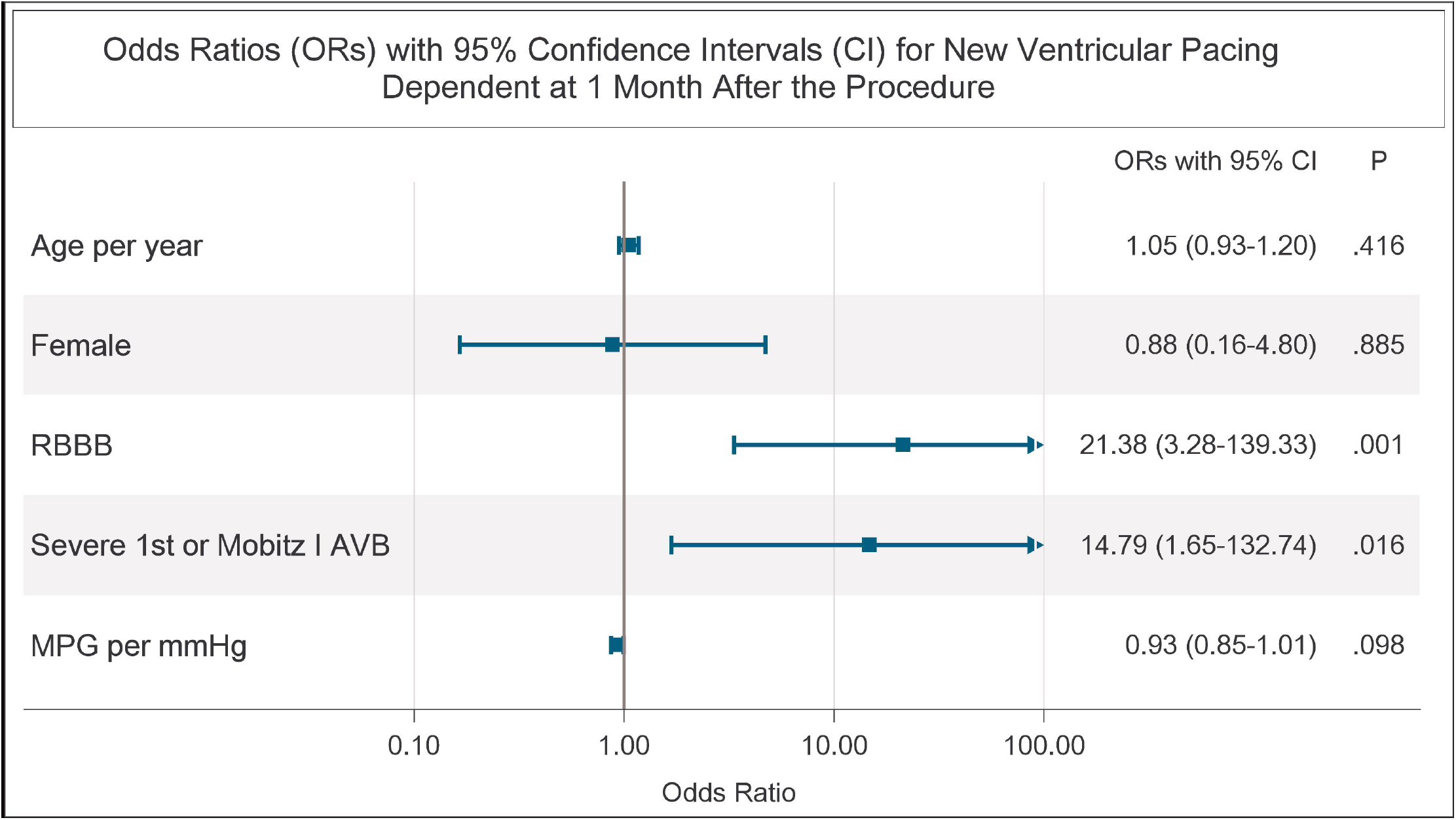
Predictors of 30-d Pacemaker Dependency by Multivariate* Binary Logistic Regression.

### Type of TAVR Devices and New VpDep

The most common type of devices was balloon-expandable device (BE; n= 86, 67.7%). The detail of all valve models was presented in Supplementary Table S2 and Table S4. The mean implantation depth was deeper in self-expanding device (SE) than the mean depth of BE (SE; 6.6 mm, SD 3.22 vs. BE; 3.5 mm, SD 1.22; P<.001). The rate of new pacemakers implanted within 1 month (Table 2 and Supplementary Table S5) was significantly lower in patients treated with BE (n=10, 12.0%) than those with SE (n=12, 32.4%; P = .008). However, the incidence of new VpDep at 1 month was similar between both groups (BE; n=6, 7.0% vs. SE; n=4, 9.8%; P=.587).

**Table 2.** Incidence of new pacemaker implantation and new ventricular pacing dependency by types of TAVR device.

| New PPM at 1 mo (n/N, %) |  | *P* | New VpDep at 1 mo |  | *P* |
| --- | --- | --- | --- | --- | --- |
| BE without prior CIED<br>(N=83) | 10/83, 12.0% | .008 | BE with prior CIED (N=3) | 1 | .587 |
|  |  |  | BE without prior CIED (N=83) | 5 |  |
|  |  |  | **Total (n/N, %)** | **6/86, 7.0%** |  |
| SE without prior CIED<br>(N=37) | 12/37, 32.4% |  | SE with prior CIED (N=4) | 2 |  |
|  |  |  | BE without prior CIED (N=37) | 2 |  |
|  |  |  | **Total (n/N, %)** | **4/41, 9.8%** |  |

Abbreviations: BE, balloon expandable device; CIED, cardiac implantable electronic device; PPM, permanent pacemaker; SE, self-expanding device; TAVR, transcatheter aortic valve replacement; VpDep, ventricular pacing dependency.

### Outcomes

At 30 days after the index procedure, 2 patients (1.6%) died, 4 (3.1%) had stroke, and 1 (0.8%) developed pacemaker infection required total removal (Supplementary Table S2). After a follow-up period of 25.8 months (SD 21.2, range 0-117 months), 18 patients (14.2%) died. New pacemaker implantation (HR 2.11; 95% CI 0.78-5.72; P = .143) and new VpDep at 30 days (HR 1.34; 95% CI 0.28-6.45; P = .720) were not associated with increased risk of death (Table 3, Supplementary Figure S2, and Figure S3).

**Table 3.** Hazard ratio and 95% confidence interval for all-cause mortality (N=127)

| **Variables** | **Hazard Ratios**<br>**(95% Confidence Interval)** | ***P*** |
| --- | --- | --- |
| Age (per year) | 1.02 (0.95-1.10) | .613 |
| Female | 0.86 (0.54-1.38) | .535 |
| Baseline LVEF (per 1%) | 0.97 (0.94-1.00) | .032 |
| STS mortality score (per 1 point) | 1.05 (1.02-1.08) | .003 |
| Old CVA | 3.29 (1.01-10.79) | .049 |
| AF or AFL | 3.99 (1.13-14.16) | .032 |
| Pre-existing RBBB | 2.90 (0.83-10.17) | .096 |
| Self-expanding TAVR device | 0.86 (0.30-2.44) | .773 |
| New pacemaker implantation within<br>30 days | 2.11 (0.78-5.72) | .143 |
| New VpDep at 1 month | 1.34 (0.28-6.45) | .720 |

Abbreviations: AF, atrial fibrillation; AFL, atrial flutter; CVA, cerebrovascular accident; LVEF, left ventricular ejection fraction; RBBB, right bundle branch block; STS, Society of Thoracic Surgeons; TAVR, transcatheter aortic valve replacement; VpDep, ventricular pacing dependency.

## Discussion

In this real-world cohort of patients underwent TAVR, 7.9% of the patients (n/N, 10/127) developed new VpDep at 1-month period after the procedure. Patients with pre-existing RBBB or Sev 1^st^/Mobitz I had more than 10-fold greater risks to develop new VpDep. The occurrence of new VpDep was not associated with higher risk of death.

TAVR has become the default therapy for severe aortic stenosis in selected patients, mostly in patients with high age and/or patients with intermediate and high surgical risks. The procedure has also now expanded to younger and low-risk patients.^10^ One of the major concerns is the need for ventricular pacing after the procedure. Long-term right ventricular pacing is known to increase risk of heart failure and all-cause mortality.^6 11^ Therefore, new VpDep developed after TAVR in patients with or without prior PPM could be prognostic. In a large cohort from Israel,^12^ high pacing burden was associated with worsened LVEF but not with excess mortality. Pacemaker dependency reportedly occurred more frequent in patients with baseline RBBB.^5 13^ Here, we showed that new VpDep occurred in less than 10% of all cases and was not associated with excess mortality after 2 years of follow-up. Underlying conduction abnormalities were strongly associated with new VpDep. Approximately 40% of patients pre-existing Sev 1^st^/Mobitz or RBBB developed new VpDep at 1 month after the procedure.

Prior studies reported a wide range of VpDep rates. In a meta-analysis,^5^ the average rate of VpDep in patients with PPM at 1 year was 47.5% with a range of 7-89%. Among these trials, the definitions of VpDep were not uniformed and the population selections were varied. In the analysis from REPRISE III trial,^14^ the cut point of 30 bpm in the absence of native rhythm was used to declare VpDep. The devices implanted in REPRISE III were all SE. The analysis reported VpDep rates of 43% and 50% at 1 month and 1 year respectively. In a trial evaluated the incidence of VpDep following Lotus valve implantation,^3^ a cut point of 40 bpm was used to define VpDep. At 30-day and 1-year, 57% and 38% were pacing dependent respectively. In a large single-center cohort including patients treated with both BE and SE devices,^13^ rates of VpDep using a cut point of 40 bpm, were 35.7% and 33.3% at 1 month and 1 year respectively. All of these trials, however, did not include patients with prior CIEDs. In the present study, we used the cut point of 30 bpm to diagnose VpDep in the absence of native rhythm. Both BE and SE devices were included. Patients with prior CIEDs who were not VpDep were also included. The VpDep rates of our analysis, calculated based on all patients with CIEDS, were approximately 34% at both 1 month and 1 year. Among patients with prior CIEDs (N=7), the rate was increased to 42.9% at both 1 month and 1 year.

Deep implantation depth has shown to be associated with conduction disturbances^15 Error! Reference source not found.^ and pacemaker dependency.^14^ The significance of Implantation depth between different devices was also varied. In addition, the optimal depth has not been consistently defined. The average implantation depths in patients with new conduction disturbances were reportedly 7.1 mm in BE^15^ and 5.2 mm in SE.^Error! Reference source not found.^ In REPRISE III trial,^14^ comparing between the Lotus valve and the CoreValve systems, one of the predictors of pacemaker dependency at 30 days was implantation depth. The mean implantation depth in REPRISE III was above 6 mm, compared to our average depth of less than 5 mm. We chose difference TAVR valve choices mainly due to anatomical suitability and found no significant differences of implantation depth between VpDep and non-VpDep groups.

The rate of PPM implantation within 1 month of procedure was 17%. The number is comparable to the rate reported in the registry that included both BE and SE devices.^2^ SE device was associated with higher rate of PPM implantation than BE device (32% vs 12%; P = .008), similar to previous data.^1 2 Error! Reference source not found.^ Our pacemaker rate was relatively high as we had relatively low threshold of pacemaker implantation. Most of the pacemaker implantation was done using index TAVR procedure visit (88% within 7 days after the index procedure). However, we showed that the incidence of new VpDep did not differ between BE and SE devices.

Our results support pacemaker interrogation and adjustment as early as 1 month. Of all patients with PPM after TAVR, approximately two-thirds did not depend on ventricular pacing at 1 month and most of them remained independent of ventricular pacing at 1 year. Appropriate pacemaker set up in this group of population would reduce unnecessary ventricular pacing and likely improve long-term outcome.

### Limitations

We recognized many limitations in our study. The sample size was relatively small. The trial design was not powered to detect effect on mortality and not allowed to assume causality. The device selection and the approach to PPM implantation was based on a single center experience. Though the maximal follow-up time was more than 9 years, the mean follow-up time was 26 months which might not be long enough to detect the consequence of VpDep.

## Conclusions

In a real-world cohort of patients underwent TAVR, the incidence of new VpDep at 1 month was 7.9% among all patients, 34.5% among patients with PPM, and 42.9% among patients with CIEDs implanted before the procedure. Pre-existing conduction abnormalities were independently associated with higher chance of new VpDep; 15 times higher for Sev 1^st^/Mobitz I and 21 times higher for RBBB. There was no excess mortality in patients who developed new VpDep at the follow-up period of 26 months.

## Data Availability

All data produced in the present study are available upon reasonable request to the authors.

