## Supplementary for "The Incidence, Predictors, and Mortality Impact of Ventricular Pacing Dependency after Transcatheter Aortic Valve Replacement: A Prospective Cohort"

### Table S1. Baseline characteristics (N=127)

| **Characters** | **Value** |
| --- | --- |
| Age, mean (SD), y | 81.8 (6.3) |
| Female, n (%) | 79 (62.2%) |
| NYHA, n (%) |  |
| - 1 | 1 (0.8%) |
| - 2 | 50 (39.4%) |
| - 3 | 69 (54.3%) |
| - 4 | 7 (5.5%) |
| Diabetes mellitus, n (%) | 90 (70.9%) |
| Hypertension, n (%) | 32 (25.2%) |
| Dyslipidemia, n (%) | 33 (26.0%) |
| Smoker, n (%) |  |
| - Never | 117 (92.1%) |
| - Current smoker | 7 (5.5%) |
| - Ex-smoker | 3 (2.4%) |
| Coronary artery disease, n (%) | 55 (43.3%) |
| Atrial fibrillation, n (%) | 105 (82.7%) |
| Bicuspid valve, n (%) | 3 (2.4%) |
| Prior CIEDs, n (%) | 7 (5.5%) |
| GFR, mean (SD), mL/min/1.73 m^2^ | 55.8 (22.60) |
| Body mass index, mean (SD), kg/m^2^ | 24.3 (4.27) |
| LVEF, mean (SD), % | 62.3 (13.7) |
| STS mortality score, mean (SD), % | 6.1 (4.5) |
| Euroscore II, mean (SD), % | 5.0 (5.2) |
| Preexisting bundle branch block, n (%) |  |
| - Intraventricular conduction delay | 1 (0.8%) |
| - Right bundle branch block | 9 (7.1%) |
| - Right bundle branch block and left anterior fascicular block | 1 (0.8%) |
| Preexisting AVB, n (%) |  |
| - 1^st^ degree AVB | 14 (11.0%) |
| - Severe 1^st^ degree AVB (PR interval >300ms) | 6 (4.7%) |
| - Mobitz I | 1 (0.8%) |
| Baseline Rhythm, n (%) |  |
| - Sinus | 119 (93.7%) |
| - Atrial fibrillation or flutter | 8 (6.3%) |

Abbreviations: AVB, atrioventricular block; CIEDs, cardiac implantable electronic devices; GFR, glomerular filtration rate; LVEF, left ventricular ejection fraction; NYHA, New York Heart Associatio; STS, Society of Thoracic Surgeons.

### Table S2. Procedural Characteristics and outcomes (N=127)

| **Characters** | **Value** |
| --- | --- |
| TAVR models, n (%) |  |
| - Absolute Neo | 17 (13.4%) |
| - EVOLUT | 6 (4.7%) |
| - Portico | 18 (14.2%) |
| - S3 | 62 (48.8%) |
| - Sapien XT | 24 (18.9%) |
| Type of TAVR devices, n (%) |  |
| - Balloon-expandable | 86 (67.7%) |
| - Self-expanding | 41 (32.3%) |
| AVA, mean (SD), cm^2^ |  |
| - Before TAVR | 0.7 (0.20) |
| - 30 days after TAVR | 1.7 (0.43) |
| Mean peak gradient, mean (SD), mmHg |  |
| - Before TAVR | 48.9 (11.90) |
| - 30 days after TVR | 10.5 (5.20) |
| Post-procedural paravalvular leak |  |
| - None | 40 (31.5%) |
| - 1+ | 65 (51.2%) |
| - 2+ | 19 (15.0%) |
| - 3+ | 3 (2.4%) |
| Implantation depth, mean (SD), mm |  |
| - At non-Coronary Cusp | 3.9 (2.45) |
| - At right Coronary Cusp | 4.8 (2.54) |
| - At left Coronary Cusp | 4.7 (2.73) |
| Procedural time, mean (SD), min | 93.5 (47.08) |
| Procedural success, n (%) | 126 (99.2%) |
| Acute complication, n (%) | 39 (30.7%) |
| - Conduction disturbances | 28 (22.0%) |
| - Major bleeding | 4 (3.1%) |
| - Cardiac tamponade | 4 (3.1%) |
| - Acute kidney injury | 4 (3.1%) |
| - Stroke | 2 (1.6%) |
| - Death | 1 (0.8%) |
| 30-day major adverse events, n (%) | 27 (21.3%) |
| PPM implantation | 22 (17.3%) |
| Stroke | 4 (3.1%) |
| Infection of pacemaker system required extraction | 1 (0.8%) |
| Death | 2 (1.6%) |
| Death at the end of follow up, n (%) | 18 (14.2%) |
| Follow-up time, mean (SD), months | 25.8 (21.21) |
| Follow-up time, range, months | 0-117 |

Abbreviations: AVA, aortic valve area; PPM, permanent pacemaker; TAVR, transcatheter aortic valve replacement.

### Table S3. Conduction disturbances and pacemaker implantation (N=127)

| **Characters** | | **Value** |
| --- | --- | --- |
| Type of new conduction disturbances, n (%) | |  |
| - Complete AVB | | 13 (10.2%) |
| - High Grade AVB | | 3 (2.4%) |
| - LBBB | | 11 (8.7%) |
| - Intraventricular conduction delay | | 2 (1.6%) |
| - 1^st^ degree AVB | | 8 (6.3%) |
| EP Study performed, n (%) | | 13 (10.2%) |
| HV interval, mean (SD), ms | | 62.9 (17.7) |
| Time to new PPM, mean (SD), days | | 47.5 (130.6) |
| New PPM, n (%) | | 25 (19.7%) |
| - Implanted ≤ 7 days after TAVR | | 22 (17.3%) |
| - Implanted >7 days to 180 days after TAVR | | 0 (0%) |
| - Implanted > 180 days after TAVR | | 3 (2.4%) |
| Indications for PPM implantation, n (%) | |  |
|  | Implanted ≤ 7 days after TAVR | Implanted > 7 days after TAVR |
| - Complete AV block | 15 (11.8%) | 3 (2.4%) |
| - New LBBB and HV interval ≥ 65ms | 5 (3.9%) | 0 (0%) |
| - High grade AVB and HV interval ≥ 65ms | 1 (0.8%) | 0 (0%) |
| - Sick sinus syndrome | 1 (0.8%) | 0 (0%) |
| Types of Pacemaker | |  |
| - Single-chamber | | 1 (0.8%) |
| - Dual-chamber | | 23 (18.1%) |
| - Cardiac resynchronization system | | 1 (0.8%) |
| New VpDep at 30 days, n (%) | | 10 (7.9%) |

Abbreviations: AVB, atrioventricular block; EP, electrophysiologic; LBBB, left bundle branch block; VpDep, ventricular pacing dependency; PPM, pacemaker; TAVR, transcatheter aortic valve replacement.

### Table S4. Pacemaker indications, parameters, and pacing dependency (n=32)

| **Patient ID** | **Time from TAVR to PPM** | **Valve** | **Indication** | **Type** | **Mode** | **LRL** | **URL** | **sAVD /pAVD** | **%VP 1mo/1yr** | **New VpDep at 1mo** | **VpDep at 1yr** |
| --- | --- | --- | --- | --- | --- | --- | --- | --- | --- | --- | --- |
| 77 | 7 years before | Portico | SSS | dPPM | DDDR | 60 | 110 | 320/350 | 36/19 | N | N |
| 74 | 4 years before | S3 | SSS | dPPM | DDD | 50 | 130 | 330/360 | 3/6 | N | N |
| 129 | 3 years before | S3 | SSS | dPPM | DDD | 50 | 110 | 250/280 | 77/100 | Y | Y |
| 110 | 1 year before | S3 | High grade AVB | dPPM | DDD | 60 | 120 | 310/340 | 54.8/59.6 | N | N |
| 137 | 6 months before | Evolut | SSS | dPPM | AAIR <=> DDDR | 60 | 110 | MVP | 34.5/17.9 | N | N |
| 63 | 2 months before | Portico | Symptomatic bifascicular block | dPPM | DDD | 50 | 100 | 200/230 | 100/99.9 | Y | Y |
| 109 | 2 months before | EVOLUT | High grade AVB | dPPM | DDD | 60 | 120 | 270/300 | 99.7/99.9 | Y | Y |
| 116 | same day | AN | CHB | dPPM | DDD | 60 | 110 | 300/330 | 2/2 | N | N |
| 141 | same day | S3 | CHB | dPPM | DDD | 60 | 110 | 200/220 | 100/100 | Y | Y |
| 21 | 1 day after | Sapien XT | CHB | dPPM | DDD | 60 | 110 | 250/280 | 8/23 | N | N |
| 119 | 1 day after | S3 | CHB | sPPM | VVI | 50 | NA | NA | 100/NA | Y | Death |
| 55 | 3 days after | S3 | CHB | dPPM | DDD | 60 | 120 | 200/220 | 0.1/16.4 | N | N |
| 64 | 3 days after | Portico | New LBBB, HV≥65 | dPPM | DDD | 60 | 120 | 240/260 | 0.1/NA | N | Death |
| 70 | 3 days after | Portico | CHB | dPPM | DDDR | 50 | 120 | 325/350 | 20/43 | N | N |
| 79 | 3 days after | Portico | New LBBB, HV≥65 | dPPM | DDD | 60 | 110 | 325/350 | 0.1/0.1 | N | N |
| 83 | 3 days after | S3 | SSS, new 1^st^ AVB | dPPM | AAIR <=> DDDR | 60 | 120 | MVP | 2.9/100 | N | Y |
| 106 | 3 days after | S3 | High grade AVB, HV≥65 | dPPM | DDD | 60 | 120 | 330/360 | 27.6/MS | N | MS |
| 134 | 3 days after | S3 | New LBBB, HV≥65 | dPPM | DDD | 60 | 100 | 300/330 | 96/0.1 | Y | N |
| 40 | 4 days after | Portico | CHB | dPPM | DDD | 60 | 120 | Search AV+ at 300 | 4/5 | N | N |
| 73 | 4 days after | S3 | New LBBB, HV≥65 | dPPM | DDI | 40 | NA | NA | 0.1/NA | N | Death |
| 80 | 4 days after | AN | CHB | dPPM | DDD | 60 | 120 | 240/260 | 90/NA | Y | Death |
| 94 | 4 days after | EVOLUT | CHB | dPPM | DDD | 60 | 120 | 300/320 | 8.4/MS | N | MS |
| 98 | 4 days after | S3 | CHB | dPPM | DDD | 60 | 120 | 270/300 | 97.7/NA | Y | Death |
| 114 | 4 days after | AN | CHB | dPPM | DDD | 50 | 120 | 200/230 | 100/99 | Y | Y |
| 135 | 4 days after | AN | CHB | dPPM | DDD | 60 | 120 | AV Search+ at 300 | 22.8/0.1 | N | N |
| 58 | 5 days after | Portico | CHB | dPPM | DDD | 60 | 120 | 200/230 | 0.2/NA | N | Death |
| 4 | 6 days after | Sapien XT | CHB | dPPM | DDD | 60 | 110 | 200/220 | 97/98.5 | Y | Y |
| 30 | 6 days after | Portico | CHB | dPPM | DDD | 60 | 120 | 240/260 | 7.7/0.1 | N | N |
| 72 | 7 days after | Portico | New LBBB, HV≥65 | dPPM | DDD | 50 | 120 | 250/275 | 0.1/0.1 | N | N |
| 81 | 7 months after | AN | CHB, HFrEF | CRT | DDD | 60 | 110 | 160/180 | 98.9^*^ | N | Y |
| 66 | 1 year after | S3 | CHB | dPPM | DDD | 50 | 100 | 250/280 | 36.6^*^ | N | NA |
| 86 | 18 months after | AN | CHB | dPPM | DDD | 60 | 100 | 180/200 | 100^*^ | N | NA |

^*^ data recorded at 1 month after implantation

Abbreviations: AN, Absolute Neo, AVB, atrioventricular block; CHB, complete heart block; CRT, cardiac resynchronization therapy; dPPM, dual-chamber pacemaker; HFrEF, heart failure with reduced ejection fraction; LBBB, left bundle branch block; LRL, lower rate limit; MS, data missing; NA, not applicable; pAVD, paced atrioventricular delay; pDep, pacemaker dependency; sAVD, sensed atrioventricular delay; sPPM, single-chamber pacemaker; SSS, sick sinus syndrome; URL, upper rate limite; %VP, ventricular pacing percentage.

### Table S5. Implantation depth, pacemaker implantation and ventricular pacing dependency by type of TAVR devices (N=127)

| **Characters** | **Type of TAVR devices** | | ***P*** |
| --- | --- | --- | --- |
|  | **Balloon-Expandable (N=86)** | **Self-Expanding (N=41)** |  |
| Implantation Depth |  |  |  |
| - At NCC, mean (SD), mm | 3.1 (1.32) | 5.7 (3.34) | <.001 |
| - At RCC, mean (SD), mm | 3.9 (1.37) | 6.8 (3.30) | <.001 |
| - At LCC, mean (SD), mm | 3.6 (1.32) | 7.1 (3.37) | <.001 |
| - Mean (SD), mm | 3.5 (1.22) | 6.6 (3.22) | <.001 |
| Prior CIEDs, n (%) | 3 (3.5%) | 4 (9.8%) | .148 |
| New Pacemaker within 1 month, n (%) | 10 (11.6%) | 12 (29.3%) | .014 |
| Indication for Pacemaker implanted within 1 month, n |  |  | .451 |
| - Complete AVB | 6 | 9 |  |
| - New LBBB and HV interval ≥ 65ms | 2 | 3 |  |
| - High grade AVB and HV interval ≥ 65ms | 1 | 0 |  |
| - Sick sinus syndrome | 1 | 0 |  |
| New VpDep at 30 days, n (%) | 6 (7.0%) | 4 (9.8%) | .587 |
| - Prior CIEDs, n | 1 | 2 |  |
| - Without prior CIEDs, n | 5 | 2 |  |
| VpDep at 1 year, n (%) | 4 (4.7%) | 4 (9.8%) | .268 |
| - Prior CIEDs, n | 1 | 2 |  |
| - Without prior CIEDs, n | 3 | 2 |  |

Abbreviations: AVB, atrioventricular block; CIEDs, cardiac implantable electronic devices; VpDep, ventricular pacing dependency.

### Table S6. Causes of Death (n=18)

| **Causes of Death** | **n (%)** |
| --- | --- |
| Aortic annulus rupture | 1 (5.6%) |
| Cardiovascular causes | 3 (16.7%) |
| Endocarditis | 1 (5.6%) |
| Malignancy | 1 (5.6%) |
| Intracranial hemorrhage | 1 (5.6%) |
| Gastrointestinal bleeding | 1 (5.6%) |
| Sepsis | 2 (11.1%) |
| Unknown | 8 (44.4%) |

### Figure S1. Timeline of Pacemaker Implantation and Incidence of Ventricular Pacing Dependency by Type of Devices

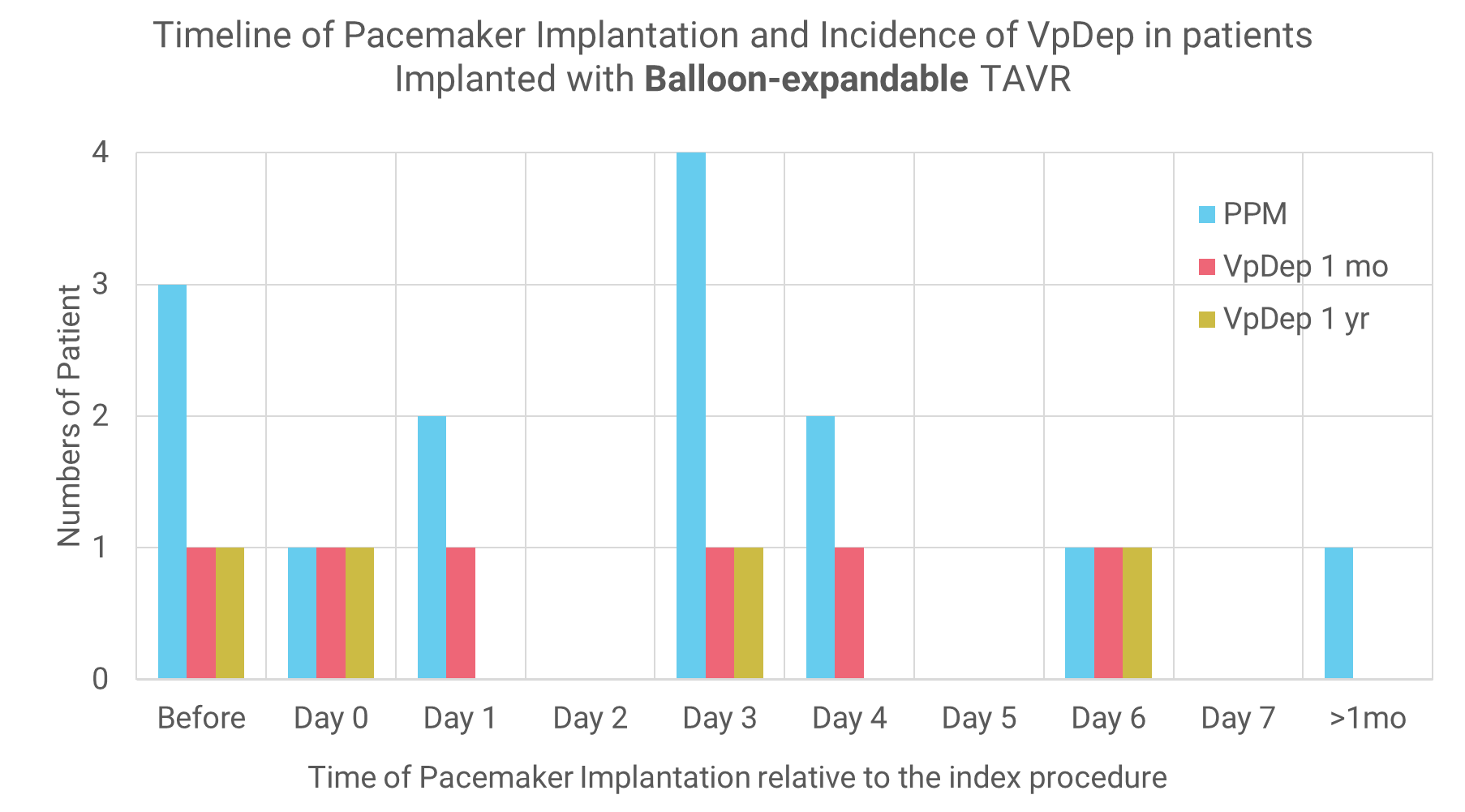

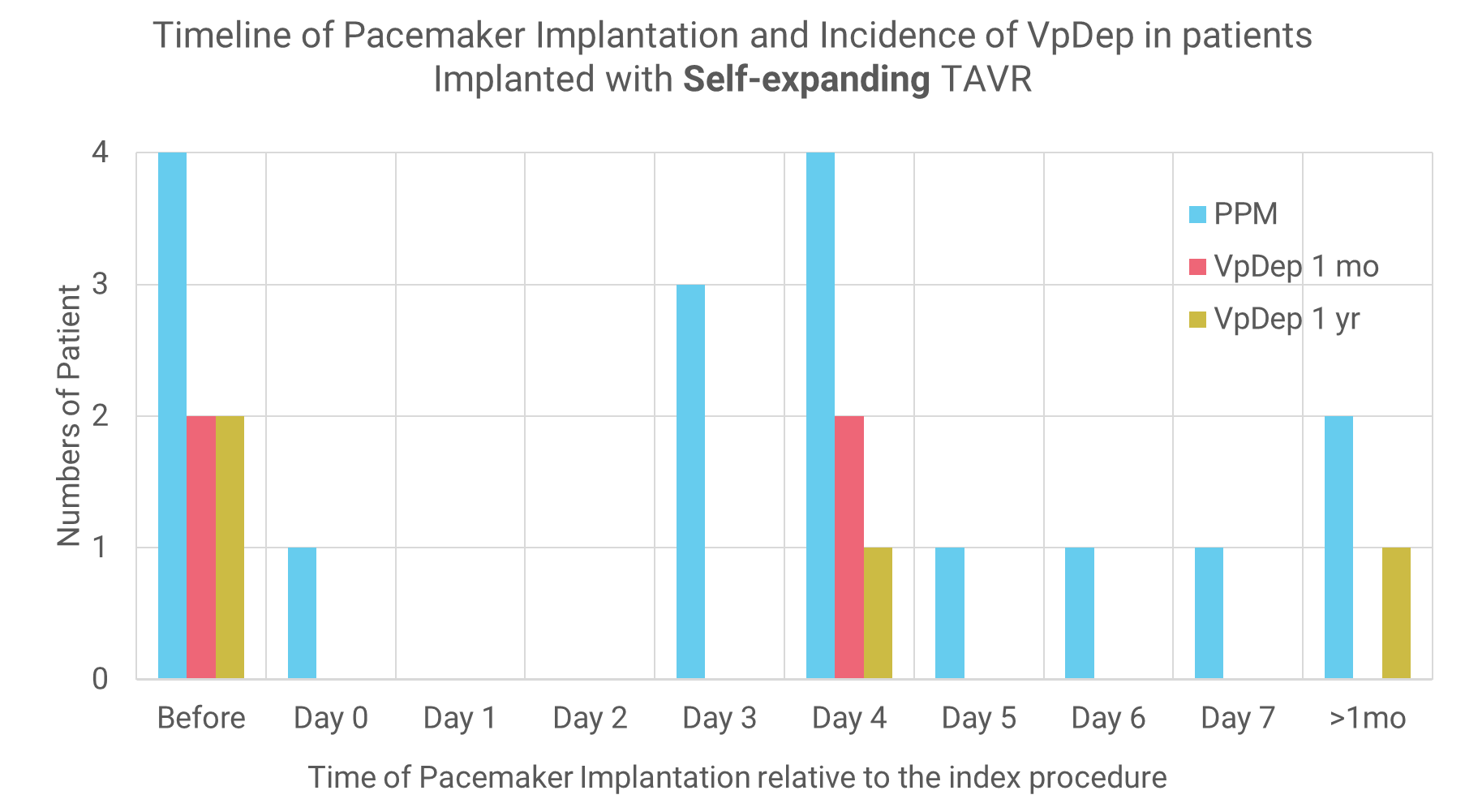

Abbreviations: TAVR, transcatheter aortic valve replacement; VpDep, ventricular pacing dependency.

### Figure S2. Kaplan-Meier survival estimates by new ventricular pacing dependency

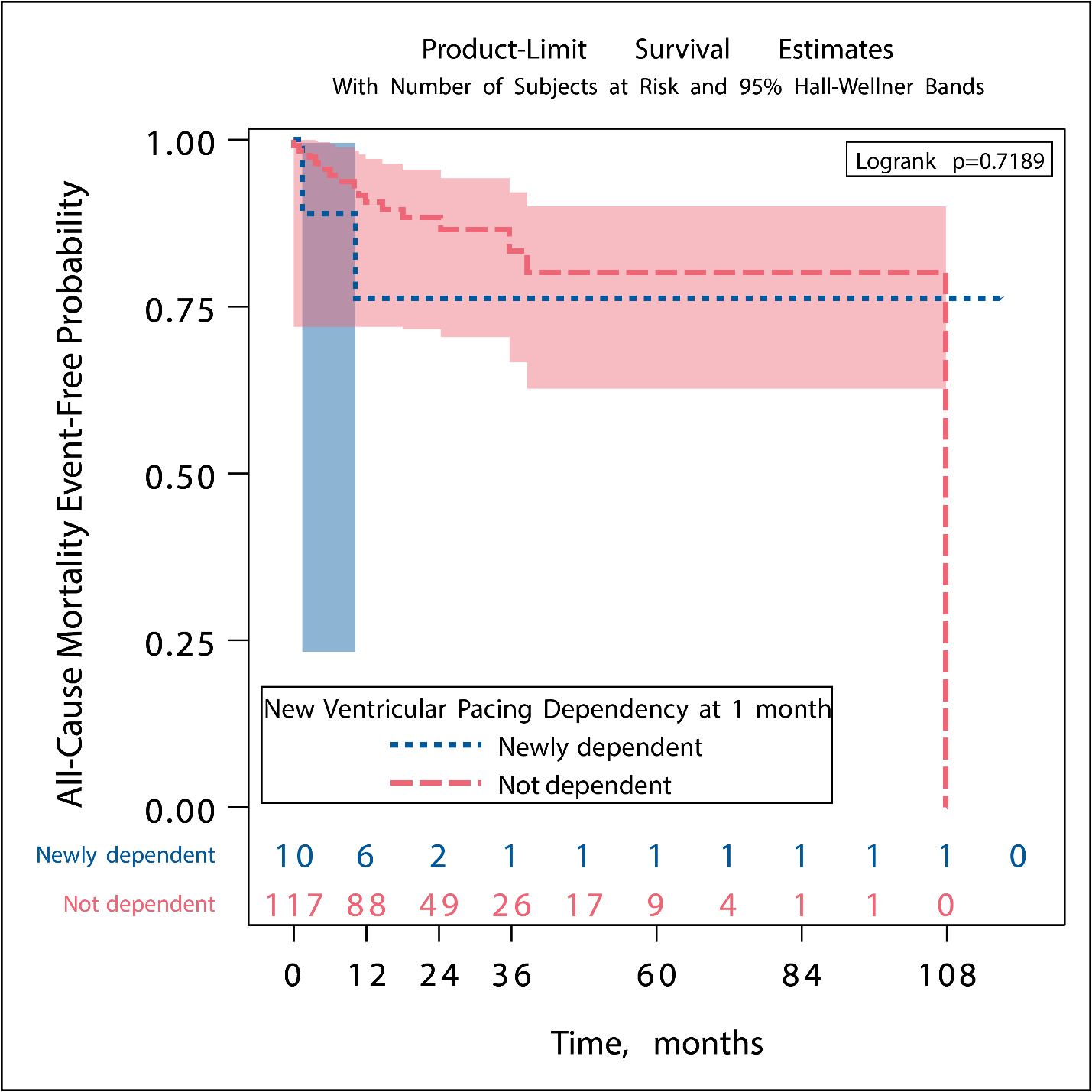

### Figure S3. Kaplan-Meier survival estimates by new permanent pacemaker implanted within 1 month after the procedure

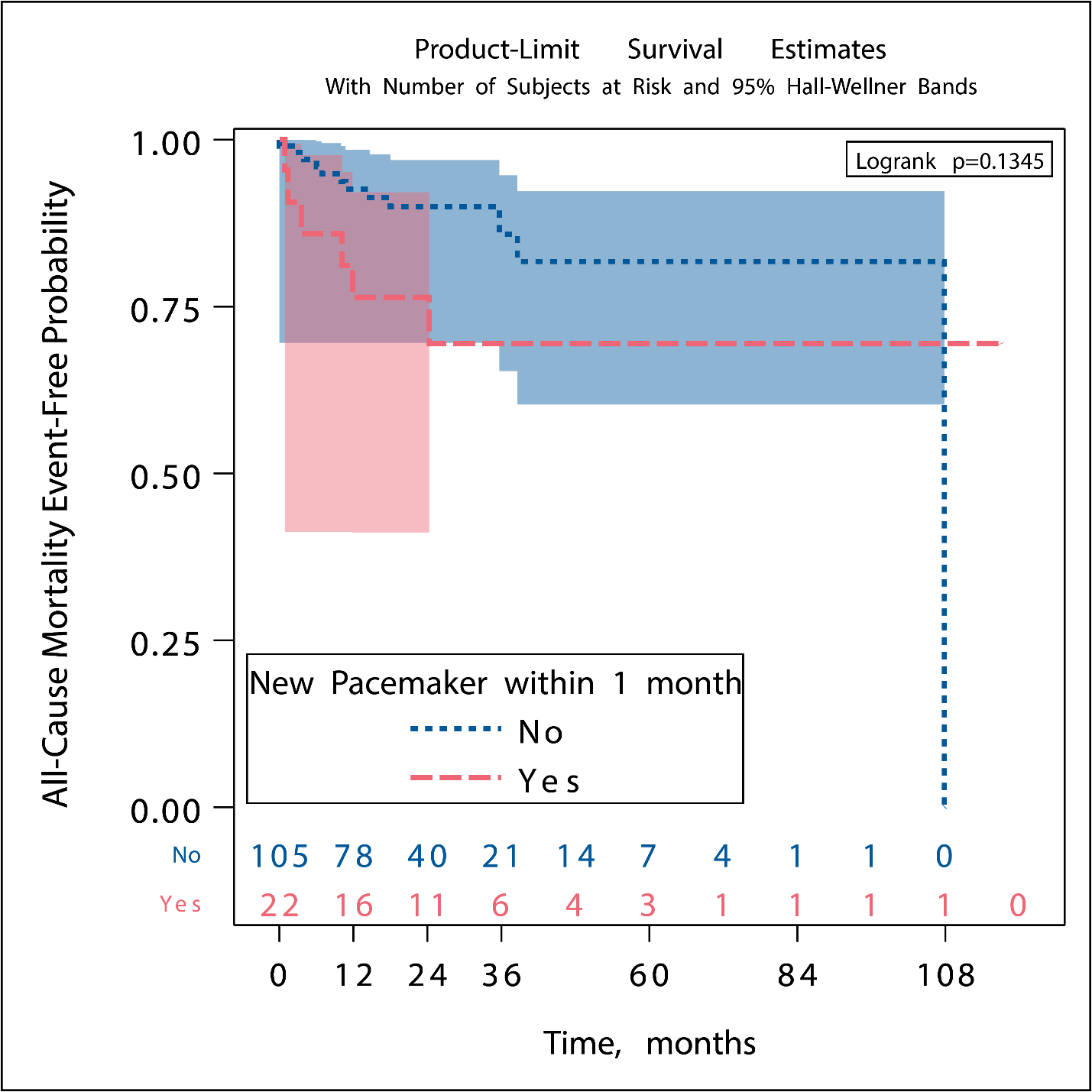
